# Assessment of heterogeneity and disease onset in the Parkinson’s Progression Markers Initiative (PPMI) cohort using the α-synuclein seed amplification assay: a cross-sectional study

**DOI:** 10.1101/2023.02.27.23286156

**Authors:** Andrew Siderowf, Luis Concha-Marambio, David-Erick Lafontant, Carly M. Farris, Yihua Ma, Paula A. Urenia, Hieu Nguyen, Roy N. Alcalay, Lana M. Chahine, Tatiana Foroud, Douglas Galasko, Karl Kieburtz, Kalpana Merchant, Brit Mollenhauer, Kathleen L. Poston, John Seibyl, Tanya Simuni, Caroline M. Tanner, Daniel Weintraub, Aleksandar Videnovic, Seung Ho Choi, Ryan Kurth, Chelsea Caspell-Garcia, Christopher S. Coffey, Mark Frasier, Luis M. A. Oliveira, Samantha J. Hutten, Todd Sherer, Kenneth Marek, Claudio Soto, the Parkinson’s Progression Markers Initiative

## Abstract

**Background:** Recent research demonstrates that α-synuclein seed amplification assays (αSyn-SAA) accurately differentiate Parkinson’s disease (PD) patients from healthy controls (HC). We used the well-characterized, multicenter Parkinson’s Progression Markers Initiative (PPMI) cohort to further assess the diagnostic performance of αSyn-SAA and to examine whether the assay identifies heterogeneity among patients and enables early identification in at-risk groups.

**Methods:** αSyn-SAA analysis of cerebrospinal fluid (CSF) was performed using previously described methods. We assessed sensitivity and specificity in PD and HC, including subgroups based on genetic and clinical features. We determined the frequency of positive αSyn-SAA results in prodromal participants (REM sleep behavior disorder and hyposmia) and non-manifesting carriers (NMCs) of genetic variants associated with PD and compared αSyn-SAA to clinical measures and other biomarkers.

**Findings:** 1,123 participants were included: 545 PD, 163 HCs, 54 participants with scans without evidence of dopaminergic deficit (SWEDDs), 51 prodromal participants, and 310 NMCs. Sensitivity and specificity for PD versus HC were 88% and 96%, respectively. Sensitivity in sporadic PD with the typical olfactory deficit was 99%. The proportion of positive αSyn-SAA was lower in subgroups including LRRK2 PD (68%) and sporadic PD patients without olfactory deficit (78%). Participants with LRRK2 variant and normal olfaction had an even lower αSyn-SAA positivity rate (35%). Among prodromal and at-risk groups, 86% of RBD and hyposmic cases had positive αSyn-SAA. 8% of NMC (either LRRK2 or GBA) were positive.

**Interpretation:** This study represents the largest analysis of αSyn-SAA for biochemical diagnosis of PD. Our results demonstrate that the assay classifies PD patients with high sensitivity and specificity, provides information about molecular heterogeneity, and detects prodromal individuals prior to diagnosis. These findings suggest a crucial role for αSyn-SAA in therapeutic development, both to identify pathologically defined subgroups of PD patients and to establish biomarker-defined at-risk cohorts.

**Funding:** PPMI – a public-private partnership – is funded by the Michael J. Fox Foundation for Parkinson’s Research and funding partners, including: PPMI is supported by a consortium of private and philanthropic parnters: Abbvie, AcureX, Aligning Science Across Parkinson’s, Amathus Therapeutics, Avid Radiopharmaceuticals, Bial Biotech, Biohaven, Biogen, BioLegend, Bristol-Myers Squibb, Calico Labs, Celgene, Cerevel, Coave, DaCapo Brainscience, 4D Pharma, Denali, Edmond J. Safra Foundation, Eli Lilly, GE Healthcare, Genentech, GlaxoSmithKline, Golub Capital, Insitro, Janssen Neuroscience, Lundbeck, Merck, Meso Scale Discovery, Neurocrine Biosciences, Prevail Therapeutics, Roche, Sanofi Genzyme, Servier, Takeda, Teva, UCB, VanquaBio, Verily, Voyager Therapeutics, and Yumanity.

**Research in Context:** *Evidence before the study:* We searched PubMed with the terms Parkinson’s disease (PD), prodromal, Non-manifest carriers, GBA, LRRK2 and real-me quaking-induced conversion (RT-QuIC), protein misfolding cyclic amplification (PMCA), and seed amplification assay (SAA) for articles published in English on or before Oct 25, 2022, in any field. This is a large and rapidly growing literature, and a number of studies were identified, including case-series of PD patients with and without genetic variants, individuals with isolated REM sleep behavior disorder (RBD), and a small number of studies of non-manifesting carriers of genetic variants (4) associated with PD.

*Added value of the study:* To our knowledge, this is the largest report of comparative data from a cohort of PD patients, healthy controls, individuals with clinical syndromes prodromal to PD (hyposmia and RBD), and non-manifest carriers of *LRRK2 G2019S* and *GBA N370S* mutations. The strengths of our data include a large sample size, robust clinical data set, a high percentage of DAT scans completed, and the ability to compare non-manifest carriers to similar aged healthy controls, which allows for inter-group comparisons and sub-group analysis. The key novel findings in this study include: 1) marked variability in rates of positive αSyn-SAA results particularly among LRRK2 variant carriers depending on olfactory performance and sex; 2) αSyn-SAA positivity in prodromal and NMCs without dopaminergic imaging abnormalities in a substantial number of cases, while the converse is less common, indicating that αSyn-SAA may be a very early indicator of synucleinopathy. We also confirmed the high diagnostic accuracy of αSyn-SAA for sporadic PD vs. HC and that αSyn-SAA is negative in the vast majority of NMC, suggesting that the presence of synuclein aggregates in CSF is not a life-long trait but rather acquired at some point relatively close to disease onset.

*Implications of all available evidence:* Our results demonstrate that the assay classifies PD patients with high sensitivity and specificity, provides information about molecular heterogeneity, and detects prodromal individuals prior to diagnosis. These findings suggest a crucial role for αSyn-SAA in therapeutic development, both to identify pathologically defined subgroups of PD patients and to establish biomarker-defined at-risk cohorts.

## Introduction

Biomarkers for Parkinson’s disease (PD) that reflect underlying pathology would improve the accuracy of early diagnosis, clarify subtypes and accelerate clinical trials.^1^ The pathological hallmark of PD is the accumulation of misfolded, aggregated α-synuclein (αSyn) in the substantia nigra and other areas of the brain.^2,3^ αSyn aggregates have also been identified in peripheral nervous system tissue.^4,5^ Recently, misfolded-protein amplification techniques, originally developed for the detection of self-propagating PrP^Sc^ in prion diseases,^6^ have been applied to detect αSyn seeds in PD and other synucleinopathies.^7,8^ These assays have been reported under the names real-me quaking-induced conversion (RT-QuIC),^9^ protein misfolding cyclic amplification (PMCA),^10^ and most recently, the consensus name, seed amplification assay (SAA).^11,12^

Prior studies have shown that αSyn seed amplification assays (αSyn-SAAs) performed on cerebrospinal fluid (CSF) distinguish PD patients from healthy controls (HC) with high sensitivity and specificity.^12,13^ Preliminary studies have also shown αSyn-SAA positive results in a high proportion of patients from at-risk groups, such as patients with isolated rapid eye movement (REM) sleep behavior disorder (RBD).^14^ A recent study showed excellent inter-laboratory agreement when samples from the same participants were run on three different assay platforms.^12^

While these and other studies have contributed to substantial progress in understanding the potential of αSyn-SAA, large-scale studies confirming and extending these results are needed. In this report, we describe αSyn-SAA results on over 1,100 participants in the Parkinson’s Progression Markers Initiative (PPMI) study, including PD patients with and without PD-associated genetic variants, healthy controls (HCs), and patients at risk for PD (either because of prodromal features or NMCs of genetic variants). The goals of this analysis were to determine assay sensitivity and specificity using a large number of samples, to leverage clinical and biomarker data within PPMI to examine clinical and genetic heterogeneity among PD patients based on αSyn-SAA status and to test the ability of αSyn-SAA to detect early signs of PD pathophysiology in at-risk individuals.

## Methods

### Study Design

This study is a cross-sectional analysis using data from the PPMI cohort. PPMI is an international, observational study with the goal of identifying clinical and biological markers of disease heterogeneity and progression in PD. This study is registered with ClinicalTrials.gov, number NCT01141023. Detailed information about inclusion criteria, informed consent, demographic data, and study design can be found at https://www.ppmi-info.org/ Data was last accessed on 12/15/2022.

### Participants

Participants in this study were recruited into one of the four PPMI cohorts, including PD patients, HCs, participants with parkinsonism but with Scans Without Evidence of Dopamine Deficiency (SWEDD), Prodromal individuals (including those with RBD or hyposmia), and NMCs of genetic variants associated with PD between 07 July 2010 and 04 July 2019. Diagnosis for each group was made by site investigators who are movement disorders specialists and confirmed by central consensus committee review. αSyn-SAA results were not available to investigators or the consensus committee at the time of diagnosis, and thus were not incorporated in classifying participants.

Sporadic PD participants (non-carriers of LRRK2 or GBA variants) were enrolled with the following criteria: within 2 years of diagnosis, not treated with PD medications at the time of enrollment, being at Hoehn and Yahr (HY) stage 1–2, had abnormal dopamine transporter (DAT) SPECT, and had two of the following: resting tremor, bradykinesia, rigidity (required to have either resting tremor or bradykinesia); or either asymmetric resting tremor or asymmetric bradykinesia. In addition, PD patients who are carriers of either the LRRK2 G2019S or the GBA N370S variants were included in this study. Inclusion criteria for genetic PD were the same as other PD participants except that genetic participants were not required to have an abnormality on DAT-SPECT, had to be within 7 years of diagnosis, and could be receiving treatment for PD.

SWEDD participants were enrolled with PD inclusion criteria, with the exception that their intial DAT-SPECT did not show evidence of decreased striatal radio-ligand uptake. HCs were age- and sex-matched healthy persons without known neurologic signs or symptoms.

Prodromal participants included individuals without a diagnosis of PD, but who had prodromal features associated with risk of PD, including RBD (confirmed by polysomnogram (PSG)) or otherwise unexplained severe hyposmia (defined as at or below the 15^th^ percentile using the University of Pennsylvania Smell Identification Test (UPSIT) (Sensonics, Philadelphia PA) in olfactory performance based on internal population norms.^15^ Enrollment of RBD and hyposmia participants was stratified to enrich for cases with abnormal DAT-SPECT. NMCs of either the LRRK2 or GBA variant noted above were included without enrichment for DAT deficit. For all participants, cohort assignment was made as described in the study protocol and confirmed by the PPMI clinical consensus review committee.

### Clinical and Pathology Assessments

All PPMI participants received a battery of clinical tests described previously.^16^ Assessments included the Movement Disorders Society (MDS) Unified Parkinson’s Disease Rating Scale (MDS-UPDRS recorded in the OFF state for treated participants), Montreal Cognitive Assessment (MoCA), rapid eye movement (REM)-sleep behavior disorder questionnaire (RBDSQ), University of Pennsylvania Smell Identification Test (UPSIT), Scales for Outcomes in Parkinson’s-Autonomic Dysfunction (SCOPA-AUT) and the 15-itme Geriatric Depression Scale (GDS). Post-mortem assessments were performed using previously described methods.^2,17^

### PPMI biomarker assessments

All participants had biosampling including blood, CSF, and urine. CSF samples were collected, stored, and shipped according to the PPMI protocol. For this study, samples collected at baseline were analyzed by αSyn-SAA (1 sample per participant). In addition to CSF αSyn-SAA assessment, CSF biomarkers included β-Amyloid 1-42 (abeta), total-tau (t-tau), phosphorylated-tau (p-tau), and quantitative total αSyn. These biomarkers were assessed using methods described previously.^18^ Plasma neurofilament light chain (NfL),^19^ and urine bis(monoacylglycerol) phosphate (BMP) were also assessed. Finally, DAT-SPECT was performed as previously described.^20^ Visual interpretation of SPECT images was used to assign patients to the PD, healthy and SWEDD cohorts. Quantitative analysis using striatal-specific binding ratios (SBR) corrected for age, and sex were used to compare DAT with clinical and biomarker data.

### αSyn seed amplification assay

The Amprion αSyn-SAA developed by Concha, Soto, and collaborators has been described previously, following a detailed protocol recently published.^21^ Briefly, the 200-μl reaction mixture included 0.3 mg/mL rec-αsyn (Amprion, cat#2020), 0.5 M NaCl (Lonza, cat#51202), 100 mM PIPES-NaOH pH 6.50 (MilliporeSigma, cat#80635), and 20% v/v CSF. Rec-αSyn was expressed with C-terminal His-tag in *E. coli* BL21(DE3) and purified using immobilized metal afinity chromatography (IMAC). All samples were analyzed with a single batch of the substrate. One 3/32" Si_3_N_4_ bead (Tsubaki Nakashima) was added per well using a house-made bead dispenser.

Beads were blocked with 1% BSA in 100 mM PIPES-NaOH pH 6.50 for 1 h and washed twice with 100mM PIPES-NaOH pH 6.50. Samples were run in 3 technical replicates within 96-well plates, using 3 FLUOstar Omega readers set to 37 °C. Each plate was shaken for 1 min every 29 min, and fluorescence was measured after each cycle for 150 h. The F_max_ (highest raw fluorescence from each well) was used in a probabilistic algorithm to determine whether each of the 3 replicates is positive or negative, and the results of the triplicate are used to determine the assay output for each sample. If all 3 replicates from a given sample were positive, the sample is deemed “positive” αSyn-SAA. If 0–1 replicates are positive, the sample is deemed “negative”. If 2 replicates are positive, the sample is deemed “inconclusive”. A second-level criterion within the algorithm compares the average maximum fluorescence (F_max_) of the 3 replicates from inconclusive samples, and samples with highly variable or low F_max_ were deemed negative. The αSyn-SAA data is available in the PPMI LONI (Laboratory of Neuroimaging) database (Amprion: project#155).

All αSyn-SAA analyses were performed blinded to participant demographic features and diagnosis. Blinding was protected by shipping samples randomized by cohort using unique specimen IDs.

### Statistical analysis

SAS 9.4 software was used for statistical analyses and figures. Sensitivity and specificity with 95% Wald confidence intervals were calculated for PD, HC, and SWEDD cohorts and selected subgroups. For total samples fewer than 40, Wilson’s method was used to calculate the 95% confidence intervals for sensitivity and specificity. Descriptive statistics at baseline including median [interquartile range (IQR)] for continuous measures and frequency (percent) for categorical measures were calculated by cohort, subgroup, and αSyn-SAA status for demographics, MDS-UPDRS scores, CSF biomarkers, UPSIT percentile, and DAT SBR. For groups with total samples fewer than 3, IQRs were not provided for continuous outcomes. Similarly, for categorical outcomes, percentages for groups with total samples fewer than 10 were not provided. Separately for each PD subgroup, the two sample Wilcoxon rank sum test was used to test for differences in distribution between αSyn-SAA positive and αSyn-SAA negative participants for age, disease duration, UPSIT percentile, and % expected DAT SBR. Moreover, to measure the association between αSyn-SAA status and sex, HY stage, race, and hyposmic status, chi-square tests or Fisher’s exact tests (when the expected count in a cell is less than 5) were used. In addition, the logistic regression model was applied to determine the relationship between the log odds of a positive αSyn-SAA status compared to a negative status for each MDS-UPDRS score and biomarker characteristic of PD subgroups. We tested models with continuous predictors with evidence of a non-linear fit for quadratic fits and compared linear and quadratic fits using BIC. As BIC did not demonstrate a preference (difference < 2), we opted to present the linear fit for simplicity. Notable odds ratio (OR) estimates and 95% Wald confidence intervals from the model can be found in the text. To account for skewness and the upper limit of detection, each CSF biomarker was dichotomized. To minimize the potential for bias from confounding factors, models involving MDS-UPDRS scores were adjusted for disease duration, while models involving CSF biomarkers were adjusted for age and sex. Due to low sample sizes, statistical analyses were not provided for GBA PD, HC, NMC, and prodromal subgroups.

## Results

### Participants Characteristics

1,123 participants were included in this analysis, including 545 patients with PD (sporadic = 373, LRRK2 G2019S variant =123, GBA N370S variant = 49), 163 HCs, 54 SWEDDs, 51 prodromal participants (hyposmia = 18, RBD = 33) and 310 NMCs (LRRK2= 159; GBA = 151). Prodromal participants were older and more likely to be male. NMCs were more likely to be female. Other demographic features are shown in Supplementary Table 1.

**Table 1:** Sensitivity of CSF αSyn-SAA for PD and subgroups and Specificity for HC and SWEDD

|  | *N* | *Specificity (95% CI)* | *Sensitivity (95% CI)* |
| --- | --- | --- | --- |
| **HC** | 163 | 96.3% (93.4, 99.2) |  |
| **HC\*** | 166 | 94.6% (91.1, 98.0) |  |
| **SWEDD** | 54 | 90.7% (83.0, 98.5) |  |
| **All PD cases** | 545 |  | 87.7% (84.9, 90.5) |
| **All PD cases\*** | 552 |  | 86.6% (83.8, 89.4) |
| **Sporadic PD** | 373 |  | 93.3% (90.8, 95.8) |
| **LRRK2 PD** | 123 |  | 67.5% (59.2, 75.8) |
| **GBA PD** | 49 |  | 95.9% (90.4, 100.0) |
| ***LRRK2 PD*** |  |  |  |
| **Males** | 65 |  | 78.5% (68.5, 88.5) |
| **Females** | 58 |  | 55.2% (42.4, 68.0) |
| **Hyposmics** | 69 |  | 89.9% (82.7, 97.0) |
| **Normosmics** | 49 |  | 34.7% (21.4, 48.0) |
| **Normosmics+Female** | 24 |  | 12.5% (4.3, 31.0) |

\*Inconclusive participants are included in this calculation and are treated as false negative for PD and false positive for HC.

The sensitivity of αSyn-SAA for detecting all PD cases, combining sporadic and genetic cases, was 87.7%. The specificity for identifying HCs was 96.3% (Table 1). A small number of cases had inconclusive αSyn-SAA results (n=10, 1.39%). Under the conservative assumption that the results for these samples were always incorrect, similar results were still obtained for sensitivity and specificity. αSyn-SAA was positive slightly more often for SWEDD participants than for HCs. This result is in keeping with the literature showing that a small number of individuals with borderline DAT imaging results have progressive parkinsonism.^22^

### PD Heterogeneity

The proportion of participants with positive αSyn-SAA varied across subgroups based on genetic and clinical features. Among genetic subgroups with manifest PD, positive αSyn-SAA was highest for GBA PD, followed by sporadic PD, and lower for LRRK2 PD (Table 2). Among clinical features, hyposmia was the most robust predictor of a positive assay (p<0.0001). Among all PD patients with hyposmia, the sensitivity of αSyn SAA was 97.1% (379/390) compared to 63.5%% (92/146) among all PD patients without olfactory dysfunction. Combining genetic and clinical features, LRRK2 PD participants with normal olfaction were more likely to be αSyn-SAA *negative* (65.3%) than αSyn-SAA *positive* (Figure 1). The likelihood of negative αSyn-SAA results was higher for female than male LRRK2 PD participants (44.8% vs. 21.5%; p=0.006). The likelihood of αSyn-SAA negative for female normosmic LRRK2 carriers was 21/24 (87.5%). αSyn-SAA results did not differ for males and females with sporadic PD.

**Table 2:**
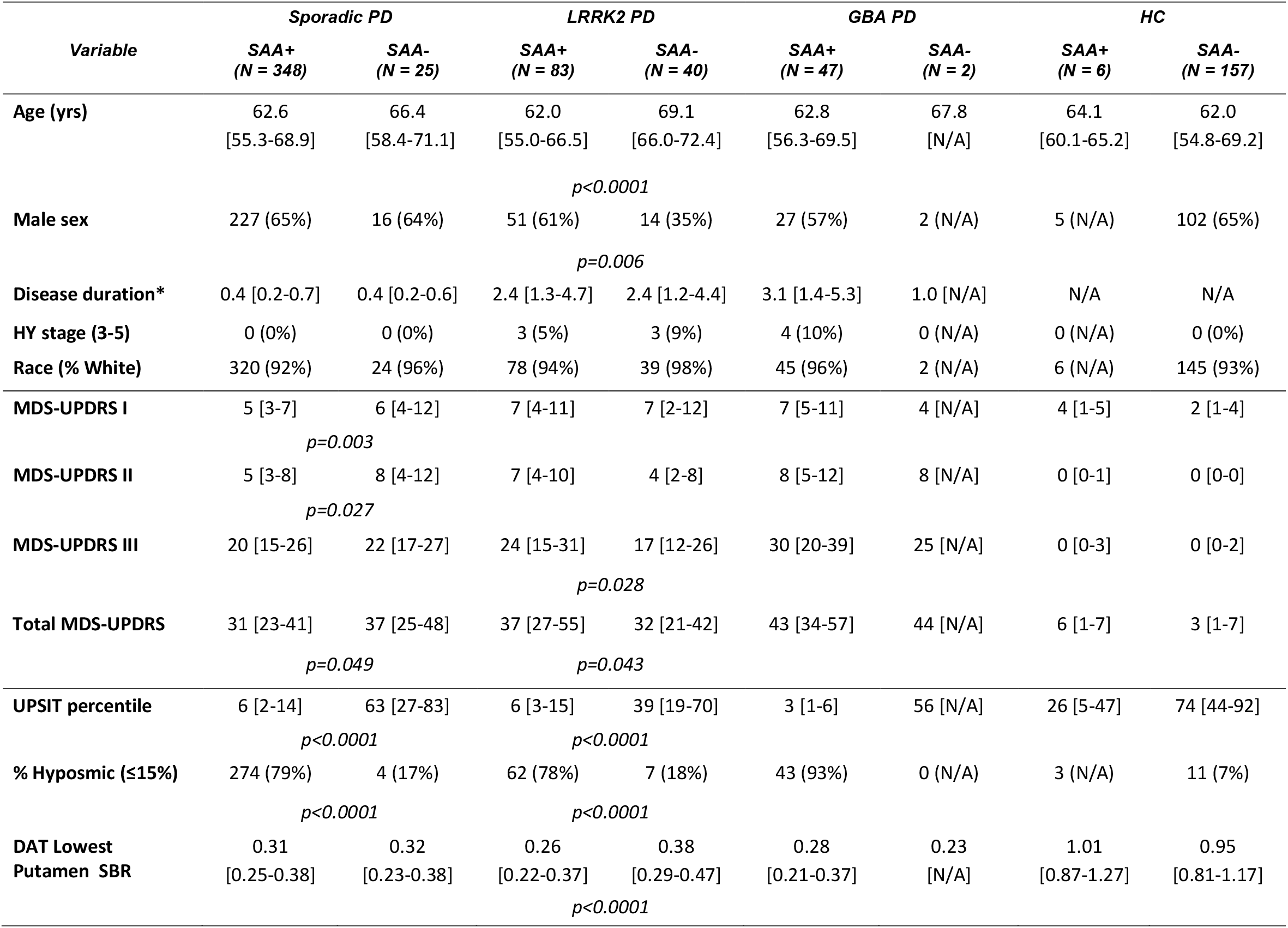
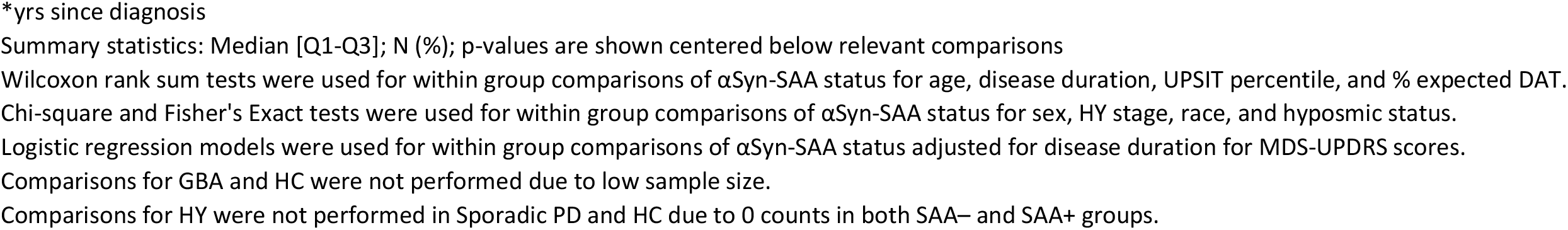
Clinical Characteristics by αSyn-SAA status for Sporadic PD, LRRK2 PD, GBA PD, and HC

**Figure 1:**
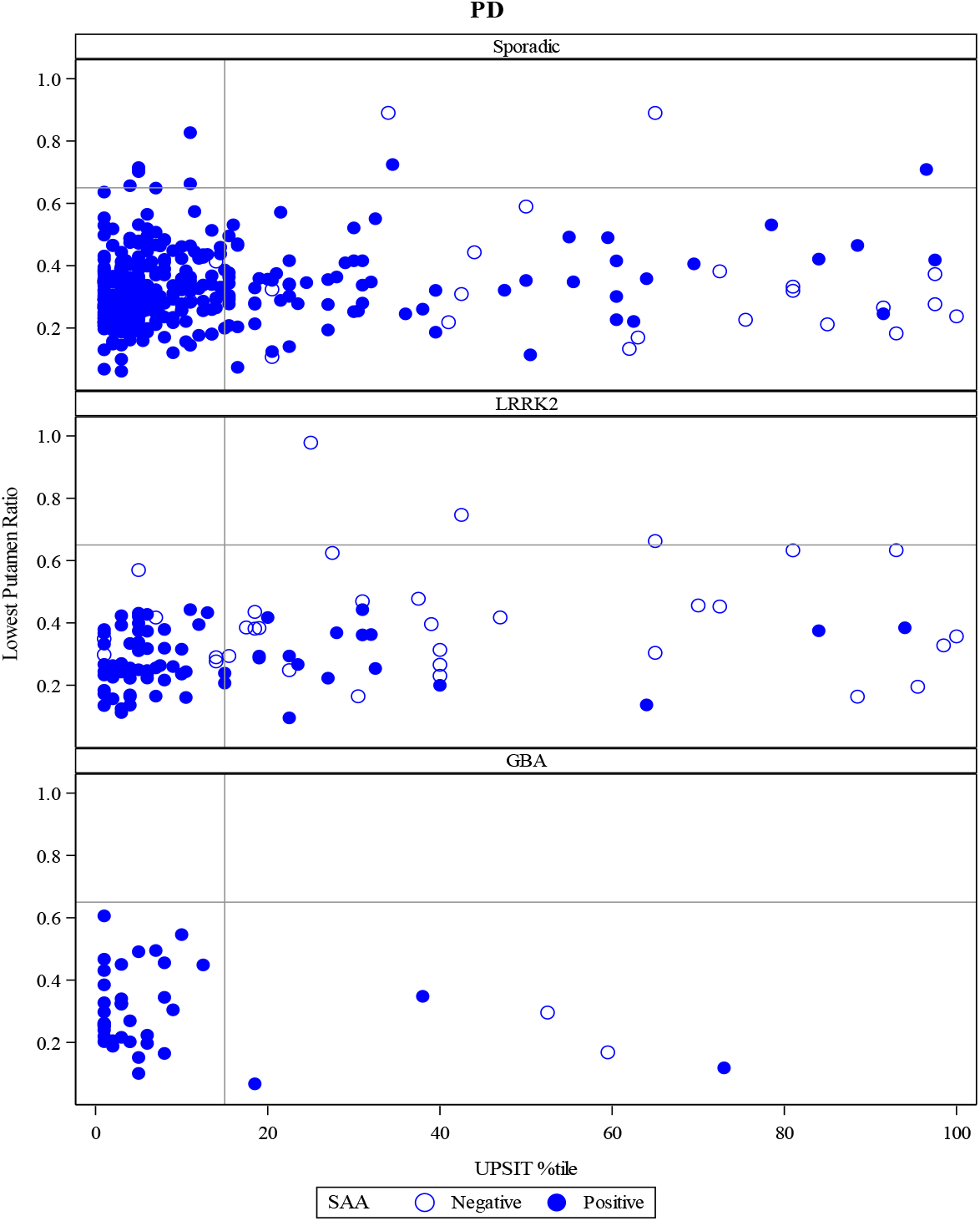
Association between DAT binding, olfaction and αSyn-SAA results among cases with manifest PD. The figure demonstrates the relationship between αSyn-SAA status and dopamine transporter imaging measured by the percent of age and sex expected lowest putamen specific binding ratio, a measure of dopamine transporter loss in the most sensitive striatal region^16^ and the UPSIT age and sex percentile of normal.^15^The horizontal line represents the DAT Lowest Putamen SBR <65% (individual below 65% are in th PD range), and the vertical line represents the age and sex adjusted UPSIT Percentile <=15% cutoff (individuals below 15% are hyposmic).

Other clinical features were significantly associated with αSyn-SAA status, but the magnitude of these associations was modest. The LRRK2 PD group with negative αSyn-SAA results had slightly less severe motor features than the αSyn-SAA positive LRRK2 PD group (p=0.028, OR= 1.053 (95% Wald CI: 1.006, 1.103)). There was a small but significant difference for sporadic PD with negative αSyn-SAA having slightly worse non-motor performance (MDS-UPDRS part I) than those with positive αSyn-SAA (p=0.003, OR=0.881 (95% Wald CI: 0.810, 0.958)). The sporadic PD group with negative αSyn-SAA results had higher MDS-UPDRS part II scores (after adjustment for disease duration) than the αSyn-SAA positive sporadic PD group (p=0.027, OR= 0.904 (95% Wald CI: (0.827, 0.989)). There were no significant associations between autonomic function, cognitive test results, depression scores or RBD scores for any groups based on αSyn-SAA status (data not shown). Likewise, there were no apparent associations with other biomarkers (Supplementary Table 2). Total BMP in urine was higher for LRRK2 than sporadic or GBA PD, but there was no difference in BMP levels among LRRK2 carriers based on αSyn-SAA status.

### Prodromal PD

The majority of RBD and hyposmic individuals had positive αSyn-SAA results including 16 of 18 hyposmics and 28 of 33 RBD (Supplementary Table 3). Among these participants, a positive αSyn-SAA result was much more likely in participants with hyposmia. For the 18 prodromal cases recruited based on smell loss, only two had negative αSyn-SAA results, and one of these was actually above the 15^th^ percentile cut-off for olfactory loss (16.5^th^ percentile). For RBD cases, all but one (of 27) αSyn-SAA positive participants were below the 15^th^ percentile while 4 of 5 αSyn-SAA negative individuals were normosmic. There were no other clinical features that were associated with a positive αSyn-SAA in prodromal participants.

Approximately 30% of the αSyn-SAA positive prodromal cases had DAT-SPECT results above the cut-off of >65% of age and sex expected uptake associated with prodromal PD,^23^ supportive of a model in which a positive αSyn-SAA result could precede abnormal DAT imaging (figure 2). A smaller number of RBD cases (3/33) had positive DAT imaging but negative αSyn-SAA results.

**Figure 2:**
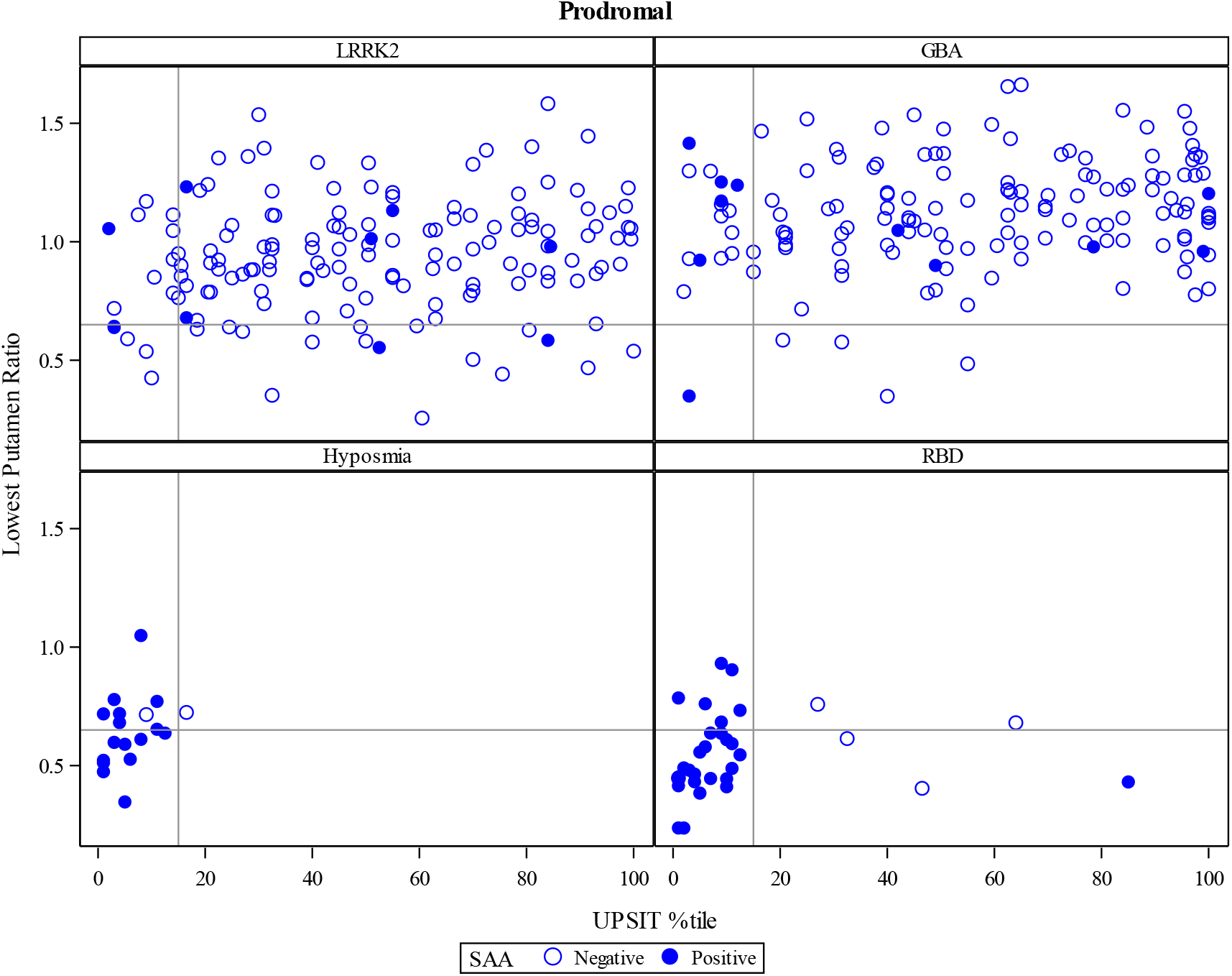
Association between DAT binding, olfaction and αSyn-SAA results among prodromal and NMC of either LRRK2 or GBA variants. The figure demonstrates the relationship between αSyn-SAA status and dopamine transporter imaging measured by the percent of age and sex expected lowest putamen specific binding ratio, a measure of dopamine transporter loss in the most sensitive striatal region^16^ and the UPSIT age and sex percentile of normal.^15^ The horizontal line represents the DAT Lowest Putamen SBR <65% (individual below 65% are in the PD range), and the vertical line represents the age and sex adjusted UPSIT Percentile <=15% cutoff (individuals below 15% are hyposmic).

While RBD and hyposmic participants were expected to be close in time to the onset of symptomatic parkinsonism because most had DAT deficit, the LRRK2 and GBA NMC were likely more distant from PD diagnosis and not required to have DAT deficit or any other risk factors. In these NMCs, 8.8% (14/159) of LRRK2 and 7.3% (11/151) of GBA NMCs had positive αSyn-SAA results.

Autopsy data were available for 15 participants, all with a clinical diagnosis of PD in life. 14 participants had typical pathology, including Lewy bodies and Lewy neurites, and were αSyn-SAA positive. The one αSyn-SAA negative case was a normosmic PD patient who also carried the LRRK2 G2019S variant. At autopsy, this patient had nigral cell loss and depigmentation but no Lewy bodies or neurites.

## Discussion

This study utilizes the comprehensive clinical and biomarker PPMI dataset to demonstrate that αSyn-SAA informs PD genetic and clinical heterogeneity and identifies at-risk individuals, possibly at a very early stage of degeneration. We confirmed that αSyn-SAA is highly accurate in differentiating PD from HCs, but observed variability among genetic sub-groups, in particular among LRRK2 PD patients. The proportion of cases with positive αSyn-SAA results was highest among patients with sporadic PD with typical olfactory deficit. By contrast, the proportion of αSyn-SAA positive results was lower in those with normal olfaction and in LRRK2 variant carriers. We found an αSyn-SAA positive gradient from almost certain in GBA PD to very likely in sporadic PD to only modestly more likely than not in LRRK2 PD. Preserved olfaction was associated with negative αSyn-SAA results across genetic sub-groups, but participants who were <u>both</u> normosmic and LRRK2 variant carriers were even more likely to be αSyn-SAA negative. This was particularly true among females, where only 12.5% of female normosmic LRRK2 carriers had a positive αSyn-SAA result.

In cases that came to post-mortem examination, all those with positive αSyn-SAA results had typical Lewy pathology, while the one case with negative αSyn-SAA, a LRRK2 carrier with preserved olfaction, had no Lewy pathology. These clinical and pathological data suggest that there may be different pathologies in αSyn-SAA positive and negative LRRK2 PD despite a similar clinical phenotype. Our results showing a lower frequency of αSyn-SAA positivity in PD patients carrying a LRRK2 variant is consistent with a prior study of 15 LRRK2 G2019S PD patients and 16 NMC.^24^ Interestingly, the proportion of αSyn-SAA negative cases that we found (approximately one third) closely mirrors the frequency of LRRK2 PD cases without Lewy pathology at autopsy reported in the literature.^25,26^ These consistent results indicate that αSyn-SAA is a marker of pathology characterized by α-Syn aggregates rather than a clinical PD phenotype, and enables ante-mortem differentiation of cases with atypical pathology.

Along with preserved olfaction, PD patients with a LRRK2 variant and a negative αSyn-SAA result were older, more likely to be female and had slightly less motor impairment. These results are consistent with a modest clinical-pathological correlation within LRRK2 PD, that can be stratified by αSyn-SAA. In addition, the excess of females among LRRK2 carriers who are αSyn-SAA negative may partially account for the observation that the expected male-to-female predominance seen in sporadic PD is not present in LRRK2 carriers.^27^

Regarding prodromal and at-risk individuals, the results of this study confirm that a majority of RBD patients with abnormal PSG and DAT deficit are αSyn-SAA positive. We have extended our assessment to individuals at-risk for PD on the basis of impaired olfaction who also had DAT deficit, and they are equally likely to be αSyn-SAA positive. Of the five (out of 33) RBD cases that were αSyn-SAA negative, three had abnormal DAT imaging, four also had normal olfactory function, possibly explained by the observation that some RBD patients progress to MSA.^28^ This disorder is associated with a less olfactory impairment than PD.^29^ Prior studies evaluating the utility of αSyn-SAA in MSA have produced varying results, depending on assay condition, and MSA has been shown to have distinct αSyn-SAA aggregation properties relative to PD.^10,30^ For this study, we used an assay that targeted detection of PD aggregates, and which has a lower sensitivity to detect glial cytoplasmic inclusions (GCIs) in MSA.

We observed several prodromal RBD and hyposmic cases with positive αSyn-SAA results but DAT imaging in the normal range. By contrast, no prodromal hyposmic participants had negative αSyn-SAA results and positive DAT imaging. Only those RBD participants potentially explained by MSA had negative αSyn-SAA and DAT binding consistent with PD. Approximately 7-10% of NMCs of either GBA or LRRK2 PD-related variants had positive αSyn-SAA results. This figure is relatively close to the lifetime penetrance of PD among GBA N370S carriers,^31^ but substantially lower than estimates of PD penetrance among LRRK2 G2019S carriers.^32,33^ Taken together, these findings are consistent with a temporal pattern of biomarker abnormalities in which there may be a prolonged period in which abnormalities in αSyn-SAA are present before changes in physiological markers such as DAT-SPECT appear, and that this pattern may be more variable in LRRK2 carriers than GBA carriers. Interestingly, among the GBA NMCs, those few individuals who are αSyn-SAA positive are also more likely to show olfactory dysfunction even in the absence of a DAT deficit. Our results provide biomarker support for a prolonged prodromal period. This concept has been proposed in the literature, lacked supporting evidence from nigral neuropathological findings or dopamine imaging studies.^34^ Subsequent studies in larger prodromal cohorts are needed to confirm this hypothesis.

The results of our study have immediate implications for clinical trial design. For LRRK2 targeted therapies, stratification based on the αSyn-SAA results may be necessary to assess therapeutic benefits. For GBA-targeted therapies it would be reasonable to exclude αSyn-SAA negative participants since GBA PD is overwhelmingly αSyn-SAA positive^35^ and, thus, a negative result would raise questions about a PD diagnosis. αSyn-SAA could also be combined with other biomarkers, such as markers of Alzheimer’s pathology to classify individuals with mixed pathology.^30^ Similarly, for αSyn targeted therapies, the possibility that αSyn-SAA negative participants will respond differently to treatment should be considered in patient selection and sample size estimates. For planned clinical trials targeting at-risk populations, αSyn-SAA results should be considered for identifying the earliest stages of synucleinopathy and those individuals likely to progress to a clinical disorder with typical Lewy pathology.

Our study has several methodological strengths. First, it uses samples from a large and well-characterized cohort consisting of several clinically relevant sub-groups. Participants were comprehensively evaluated with a battery of clinical evaluations of motor and non-motor features and biomarkers, including DAT imaging and fluid biomarkers, permitting interrogation of αSyn-SAA results against these measures. The number of PD participants and concurrent, matched controls and the inclusion of prodromal cohorts in PPMI allowed for sub-group analysis and extension of prior results to new populations. Second, αSyn-SAA analysis was performed in a blinded fashion using a robust, validated assay platform. Third, the participants were diagnosed by expert neurologists based on standardized criteria within the PPMI study.

A number of limitations of this study should be acknowledged, First, there was no a priori hypothesis or sample size estimation. Rather, we took advantage of the large and readily available PPMI cohort. Based on the diagnostic performance of the assay in prior studies,^7^ we expect a high power. We feel that this is a reasonable expectation given that this is the largest cohort that has ever been investigated in this manner. Second, the non-parametric Wilcoxon rank sum test was used to compare αSyn-SAA status in PD participants for some outcomes due to the low number of samples in some groups and skewed nature of the data, but additional samples would enable the use of more powerful methods. Third, the analyses presented in this report are all cross-sectional. Since PPMI has clinical and biological samples collected over time, a report on longitudinal data, including motor and non-motor features, will be an important topic for future analysis and may identify other clinical correlates. Fourth, the number of prodromal (hyposmia and RBD) cases with normal DAT imaging is relatively small, which prevents us from drawing definitive conclusions about the temporal ordering of αSyn-SAA abnormalities and those on DAT imaging. Fifth, several unanswered questions remain regarding genetic forms of PD. This cohort consisted of only G2019S LRRK2 carriers and N370S GBA carriers. αSyn-SAA results in patients with other LRRK2 or GBA variants, as well as variants in PD-associated genes such as PRKN and PINK1 could not be assessed.

Again, longitudinal studies and those in additional PPMI participants focusing on at-risk individuals, who are currently being recruited, may address these issues.

In summary, this study extends our understanding of the usefulness of αSyn-SAA for *in vivo* molecular assessment of PD. We show in a large, deeply phenotyped cohort that αSyn-SAA is highly accurate in typical PD, but results vary depending the presence of the LRRK2 G2019S variant as well as clinical features, particularly hyposmia. Within LRRK2 variant carriers, there are also differences in age, gender and motor performance that associate with αSyn-SAA status. Another key finding is that prodromal and NMCs, especially carriers of the GBA N370S variant, have evidence of abnormal α-synuclein aggregation prior to any other detectable clinical or biomarker change, including alterations in DAT imaging. One implication of this result is that the prodromal period in PD may be longer than had been projected previously and may start before there is loss of dopaminergic integrity, at least in some individuals. Taken together, these findings have immediate implications for clinical trial design, both to identify pathologically defined subgroups of PD patients and to establish biomarker-defined at-risk cohorts. Longitudinal research is needed to investigate the prognostic value of αSyn-SAA and whether changes in quantitative measures of αSyn aggregation indicate progressive pathology over time.

**Table S1.** Demographics and Clinical Features

| **Variable**<br>**Mean (SD)** | **Parkinson's**<br>**Disease**<br>**(N=545)** | **Healthy Control**<br>**(N=163)** | **SWEDD**<br>**(N=54)** | **Prodromal**<br>**(N=51)** | **NMC**<br>**(N=310)** |
| --- | --- | --- | --- | --- | --- |
| **Age (yrs)** | 63.4 [56.3-69.1] | 62.6 [55.0-69.2] | 63.3 [52.4-68.5] | 67.7 [65.9-73.2] | 61.7 [56.6-66.8] |
| **Male sex** | 337 (62%) | 107 (66%) | 32 (59%) | 40 (78%) | 134 (43%) |
| **Disease duration (yrs since diagnosis)** | 0.6 [0.3-1.8] | N/A | 0.3 [0.2-0.9] | N/A | N/A |
| **HY stage (3-5)** | 10 (2%) | 0 (0%) | 1 (2%) | 0 (0%) | 0 (0%) |
| **Race** |  |  |  |  |  |
| White | 508 (93%) | 151 (93%) | 51 (94%) | 48 (96%) | 299 (98%) |
| Non-white | 36 (7%) | 11 (7%) | 3 (6%) | 2 (4%) | 6 (2%) |
| American Indian or Alaska Native | 1 (0%) | 0 (0%) | 0 (0%) | 0 (0%) | 0 (0%) |
| Asian (including Native Hawaiian<br>OR other Pacific Islander) | 9 (2%) | 0 (0%) | 1 (2%) | 0 (0%) | 0 (0%) |
| Black | 6 (1%) | 8 (5%) | 1 (2%) | 1 (2%) | 0 (0%) |
| Multiracial | 17 (3%) | 2 (1%) | 0 (0%) | 0 (0%) | 4 (1%) |
| Not reported | 3 (1%) | 1 (1%) | 1 (2%) | 1 (2%) | 2 (1%) |
| **Hispanic or Latino** | 31 (6%) | 3 (2%) | 2 (4%) | 18 (35%) | 15 (5%) |
| **UPSIT percentile** | 6 [3-17] | 74 [40-92] | 44 [21-77] | 7 [3-11] | 51 [29-79] |
| % Hyposmic (<=15%) | 390 (73%) | 14 (9%) | 10 (19%) | 44 (88%) | 40 (13%) |
| **DAT Lowest Putamen SBR** | 0.30 [0.24-0.38] | 0.95 [0.81-1.17] | 0.95 [0.81-1.13] | 0.59 [0.45-0.71] | 1.04 [0.88-1.20] |
| **Positive αSyn-SAA** | 478 (88%) | 6 (4%) | 5 (9%) | 44 (86%) | 25 (8%) |
| **% LRRK2+** | 123 (23%) | 0 (0%) | 0 (0%) | N/A | 159 (51%) |
| **% GBA+** | 49 (9%) | 0 (0%) | 0 (0%) | N/A | 151 (49%) |

Summary statistics: Median [Q1-Q3]; N (%)
Statistical analyses comparing cohorts were not performed.

**Table S2:** Biomarker characteristics by αSyn-SAA status for Sporadic PD, LRRK2 PD, GBA PD, and HC

| Variable | Sporadic PD |  | LRRK2 PD |  | GBA PD |  | HC |  |
| --- | --- | --- | --- | --- | --- | --- | --- | --- |
|  | SAA+<br>(N = 348) | SAA-<br>(N = 25) | SAA+<br>(N = 83) | SAA-<br>(N = 40) | SAA+<br>(N = 47) | SAA-<br>(N = 2) | SAA+<br>(N = 6) | SAA-<br>(N = 157) |
| Mean striatum SBR | 1.35 [1.14-1.62] | 1.26 [1.04-1.84] | 1.24 [0.97-1.54] | 1.39 [1.06-1.49] | 1.16 [0.92-1.56] | 0.80 [N/A] | 2.45 [2.09-3.16] | 2.44 [2.19-2.90] |
| CSF abeta (< 683) | 104 (32%) | 8 (35%) | 27 (33%) | 13 (34%) | 23 (50%) | 0 (N/A) | 0 (N/A) | 41 (27%) |
| CSF tau (< 266) | 303 (93%) | 22 (92%) | 77 (93%) | 32 (82%) | 44 (96%) | 2 (N/A) | 5 (N/A) | 135 (87%) |
| CSF αsyn (< 1000) | 71 (22%) | 3 (13%) | 11 (24%) | 2 (10%) | 4 (50%) | N/A | 1 (N/A) | 24 (15%) |
| Serum NfL | 11.50 [8.30-15.80] | 13.85 [11.60-19.10] | 10.70 [8.77-14.95] | 18.60 [10.50-30.00] | 12.90 [9.58-17.50] | 26.70 [N/A] | 12.45 [9.14-16.10] | 10.30 [7.09-14.80] |
| Total di-18:1 BMP | 3.7 [2.1-6.8] | 2.9 [1.8-5.4] | 10.9 [7.7-20.7] | 15.0 [6.5-29.1] | 5.9 [2.4-9.5] | 12.9 [N/A] | 5.6 [2.7-11.7] | 3.5 [1.9-6.4] |

Summary statistics: Median [Q1-Q3]; N (%)
Logistic regression models were used for within group comparisons of SAA αSyn-SAA status adjusted for age and sex.
Comparisons for GBA and HC were not performed due to low sample size.

**Table S3:** Clinical Characteristics for NMC, Hyposmia, and RBD

| *Variable* | *LRRK2* |  | *GBA* |  | *Hyposmia* |  | *RBD* |  |
| --- | --- | --- | --- | --- | --- | --- | --- | --- |
|  | *SAA+*<br>*(N = 14)* | *SAA-*<br>*(N = 145)* | *SAA+*<br>*(N = 11)* | *SAA-*<br>*(N = 140)* | *SAA+*<br>*(N = 16)* | *SAA-*<br>*(N = 2)* | *SAA+*<br>*(N = 28)* | *SAA-*<br>*(N = 5)* |
| **Age (yrs)** | 63.3 [57.7-76.4] | 61.5 [56.0-67.1] | 61.1 [60.5-70.8] | 62.1 [57.3-66.5] | 66.4 [63.4-70.1] | 78.2 [N/A] | 69.7 [66.3-73.1] | 70.8 [62.4-77.0] |
| **Male sex** | 7 (50%) | 59 (41%) | 5 (45%) | 63 (45%) | 10 (63%) | 2 (N/A) | 23 (82%) | 5 (N/A) |
| **Race (% White)** | 12 (92%) | 141 (99%) | 11 (100%) | 135 (97%) | 15 (94%) | 2 (N/A) | 27 (100%) | 4 (N/A) |
| **MDS-UPDRS I** | 3 [2-6] | 4 [2-6] | 3 [1-9] | 4 [2-7] | 4 [2-7] | 6 [N/A] | 9 [4-11] | 6 [4-7] |
| **MDS-UPDRS II** | 1 [0-2] | 0 [0-1] | 0 [0-2] | 0 [0-1] | 2 [0-3] | 2 [N/A] | 2 [0-5] | 1 [1-3] |
| **MDS-UPDRS III** | 2 [1-4] | 2 [0-4] | 0 [0-4] | 1 [0-4] | 1 [0-5] | 5 [N/A] | 3 [2-6] | 5 [3-8] |
| **Total MDS-UPDRS** | 7 [4-12] | 6 [4-12] | 3 [2-24] | 7 [3-13] | 8 [4-17] | 13 [N/A] | 14 [8-19] | 14 [8-18] |
| **UPSIT percentile** | 17 [7-55] | 50 [28-77] | 12 [5-79] | 57 [32-84] | 5 [2-8] | 13 [N/A] | 7 [3-10] | 33 [27-47] |
| % Hyposmic (<=15%) | 6 (43%) | 15 (10%) | 6 (55%) | 13 (9%) | 16 (100%) | 1 (N/A) | 26 (96%) | 1 (N/A) |
| **DAT Lowest Putamen SBR** | 0.83 [0.64-1.06] | 0.94 [0.82-1.10] | 1.05 [0.92-1.24] | 1.13 [0.99-1.28] | 0.62 [0.52-0.72] | 0.72 [N/A] | 0.52 [0.44-0.64] | 0.61 [0.45-0.68] |

Summary statistics: Median [Q1-Q3]; N (%)
Statistical analyses were not provided due to small numbers in some groups.

## Supporting information

Author Contributions

PPMI Author List for Publications

PPMI Data and Funding Statement

## Data Availability

All data produced are available online at https://www.ppmi-info.org/access-data-specimens/download-data

