## Supplementary material for "Assessment of heterogeneity and disease onset in the Parkinson’s Progression Markers Initiative (PPMI) cohort using the α-synuclein seed amplification assay: a cross-sectional study": Author Contributions

| | Author | Highest degree | Full professor | Roles |
|---|---|---|---|---|
| 1 | Andrew Siderowf | MD | Yes | conceptualization, data curation, formal analysis, investigation, methodology, supervision, validation, writing – original draft, and writing– review & editing. |
| 1 | Luis Concha-Marambio | PhD | | conceptualization, data curation, formal analysis, investigation, methodology, validation, writing – original draft, and writing– review & editing. |
| 3 | David-Erick Lafontant | MS | | conceptualization, data curation, formal analysis, methodology, validation, and writing– review & editing. |
| 4 | Carly M. Farris | MS | | data curation, formal analysis, methodology, validation, and writing– review & editing. |
| 5 | Yihua Ma | MS | | data curation, formal analysis, methodology, validation, and writing– review & editing. |
| 6 | Paula A. Urenia | BA | | data curation, formal analysis, methodology, validation, and writing– review & editing. |
| 7 | Hieu Nguyen | MS | | data curation, formal analysis, methodology, validation, and writing– review & editing. |
| 8 | Roy N. Alcalay | MD | Yes | conceptualization, investigation, methodology, supervision, and writing– review & editing. |
| 9 | Lana M. Chahine | MD | | conceptualization, investigation, methodology, supervision, and writing– review & editing. |
| 10 | Tatiana Foroud | PhD | Yes | conceptualization, investigation, methodology, supervision, and writing– review & editing. |
| 11 | Douglas Galasko | MD | Yes | conceptualization, investigation, methodology, supervision, and writing– review & editing. |
| 12 | Karl Kieburtz | MD | Yes | conceptualization, investigation, methodology, supervision, and writing– review & editing. |
| 13 | Kalpana Merchant | PhD | Yes | conceptualization, investigation, methodology, supervision, and writing– review & editing. |
| 14 | Brit Mollenhauer | MD | Yes | conceptualization, investigation, methodology, supervision, and writing– review & editing. |
| 15 | Kathleen L. Poston | MD | Yes | conceptualization, investigation, methodology, supervision, and writing– review & editing. |
| 16 | John Seibyl | MD | | conceptualization, investigation, methodology, supervision, and writing– review & editing. |
| 17 | Tanya Simuni | MD | Yes | conceptualization, investigation, methodology, supervision, and writing– review & editing. |

| | | | | |
|---|---|---|---|---|
| 18 | Caroline M. Tanner | MD | yes | conceptualization, investigation, methodology, supervision, and writing–review & editing. |
| 19 | Daniel Weintraub | MD | yes | conceptualization, investigation, methodology, supervision, and writing–review & editing. |
| 20 | Aleksandar Videnovic | MD | | conceptualization, investigation, methodology, supervision, and writing–review & editing. |
| 21 | Seung Ho Choi | MS | | data curation, formal analysis, investigation, methodology, validation, writing – original draft, and writing– review & editing. |
| 22 | Ryan Kurth | MS | | data curation, formal analysis, investigation, methodology, validation, writing – original draft, and writing– review & editing. |
| 23 | Chelsea Caspell-Garcia | MS | | data curation, formal analysis, investigation, methodology, validation, writing – original draft, and writing– review & editing. |
| 24 | Christopher S. Coffey | PhD | yes | conceptualization, data curation, formal analysis, investigation, methodology, project administration, supervision, and writing–review & editing. |
| 25 | Mark Frasier | PhD | | conceptualization, funding acquisition, investigation, methodology, project administration, resources, and writing–review & editing. |
| 26 | Luis M. A. Oliveira | PhD | | conceptualization, funding acquisition, investigation, methodology, project administration, resources, and writing–review & editing. |
| 27 | Samantha J. Hutten | PhD | | conceptualization, funding acquisition, investigation, methodology, project administration, resources, and writing–review & editing. |
| 28 | Todd Sherer | PhD | | conceptualization, funding acquisition, investigation, methodology, project administration, resources, and writing–review & editing. |
| 29 | Kenneth Marek | MD | | conceptualization, data curation, formal analysis, funding acquisition, investigation, methodology, project administration, resources, supervision, and writing– review & editing. |
| 30 | Claudio Soto | PhD | yes | conceptualization, investigation, methodology, project administration, supervision, validation, and writing– review & editing. |
