## Supplementary material for "Assessment of heterogeneity and disease onset in the Parkinson’s Progression Markers Initiative (PPMI) cohort using the α-synuclein seed amplification assay: a cross-sectional study": PPMI Author List for Publications

### PPMI STUDY TEAMS/CORES/COLLABORATORS FOR PUBLICATIONS

**Executive Steering Committee:**
Kenneth Marek, MD[1] (Principal Investigator); Caroline Tanner, MD, PhD[9]; Tanya Simuni, MD[3]; Andrew Siderowf, MD, MSCE[12]; Douglas Galasko, MD[27]; Lana Chahine, MD[41]; Christopher Coffey, PhD[4]; Kalpana Merchant, PhD[61]; Kathleen Poston, MD[40]; Roseanne Dobkin, PhD[43]; Tatiana Foroud, PhD[15]; Brit Mollenhauer, MD[8]; Dan Weintraub, MD[12]; Ethan Brown, MD[9]; Karl Kieburtz, MD, MPH[23]

**Steering Committee:**
Duygu Tosun-Turgut, PhD[9]; Werner Poewe, MD[7]; Susan Bressman, MD[14]; Jan Hammer[11]; Raymond James, RN[22]; Ekemini Riley, PhD[42]; John Seibyl, MD[1]; Leslie Shaw, PhD[12]; David Standaert, MD, PhD[18]; Sneha Mantri, MD, MS[62]; Nabila Dahodwala, MD[12]; Michael Schwarzschild[47]; Connie Marras[45]; Hubert Fernandez, MD[25]; Ira Shoulson, MD[23]; Helen Rowbotham[2]; Lucy Norcliffe-Kaufmann[2]; Claudia Trenkwalder, MD[8]

**Michael J. Fox Foundation (Sponsor):** Todd Sherer, PhD; Sohini Chowdhury; Mark Frasier, PhD; Jamie Eberling, PhD; Katie Kopil, PhD; Alyssa O'Grady; James Gibaldi, MSc; Maggie McGuire Kuhl; Leslie Kirsch, EdD

**Study Cores, Committees and Related Studies:** ***(Include as applicable to the paper)***
*Project Management Core:* Emily Flagg[1]
*Site Management Core:* Tanya Simuni, MD[3]; Bridget McMahon[1]
Strategy and Technical Operations: Craig Stanley[1]; Kim Fabrizio[1]
*Data Management Core:* Dixie Ecklund, MBA, MSN[4]; Trevis Huff[4]; Richard Peters[4]; Janel Fedler[4]
*Screening Core:* Tatiana Foroud, PhD[15]; Laura Heathers[15]; Christopher Hobbick[15]; Gena Antonopoulos[15]
*Imaging Core:* John Seibyl, MD[1]; Kathleen Poston, MD[40]
*Statistics Core*: Christopher Coffey, PhD[4]; Chelsea Caspell[4]; Michael Brumm, MS[4]
*Bioinformatics Core*: Arthur Toga, PhD[10]; Karen Crawford[10]
*Biorepository Core:* Tatiana Foroud, PhD[15]; Jan Hammer[11]
*Biologics Review Committee*: Brit Mollenhauer[8]; Doug Galasko[27]; Kalpana Merchant[61]
*Genetics Core:* Andrew Singleton, PhD[13]
*Pathology Core:* Tatiana Foroud, PhD[15]; Thomas Montine, MD, PhD[40]
*Found:* Caroline Tanner, MD PhD[9]
*PPMI Online:* Carlie Tanner, MD PhD[9]; Ethan Brown[9]; Lana Chahine[41]; Roseann Dobkin[43]; Monica Korell[9]

**Site Investigators:**
Ruth Schneider, MD[23]; Kelvin Chou, MD[44]; David Russell, MD, PhD[1]; Stewart Factor, DO[16]; Penelope Hogarth, MD[17]; Robert Hauser, MD, MBA[19]; Nabila Dahodwala, MD, MSc[12]; Marie H Saint-Hilaire, MD, FRCPC, FAAN[22]; David Shprecher, DO[24]; Hubert Fernandez, MD[25]; Kathrin Brockmann, MD[26]; Yen Tai, MD, PhD[29]; Paolo Barone, MD, PhD[30]; Stuart Isaacson, MD[31]; Alberto Espay, MD, MSc, FAAN, FANA[32]; Maria Jose Martí, MD, PhD[34]; Eduardo Tolosa MD, PhD[34]; Shu-Ching Hu, MD, PhD[21]; Douglas Galasko, MD[27]; Emile Moukheiber, MD[28]; Jean-Christophe Corvol, MD[39]; Nir Giladi, MD[36]; Javier Ruiz Martinez, MD, PhD[35]; Jan O. Aasly, MD[37]; Leonidas Stefanis, MD, PhD[38]; Karen Marder, MD MPH[39]; Arjun Tarakad, MD[20]; Connie Marras, MD, PhD, FRCP(C)[45]; Tiago Mestre, MD, PhD[46]; Aleksandar Videnovic, MD, MSc[47]; Rajesh Pahwa, MD[48]; Mark Lew, MD[49]; Holly Shill, MD[50]; Amy Amara, MD, PhD[18]; Charles Adler, MD, PhD[51]; Caroline Tanner, MD, PhD[9]; Susan Bressman, MD[14]; Tanya Simuni, MD[3]; Maureen Leehey, MD[52]; Giulietta Riboldi, MD[53]; Nikolaus McFarland, MD, PhD, FAAN[54]; Lana Chahine, MD[41]; Ron Postuma, MD, FRCPC[55]; Brit Mollenhauer, MD[8]; Werner Poewe, MD[7]; Zoltan Mari, MD[56]; Nicola Pavese, MD, PhD[57]; Michele Hu, MD, PhD[58]; Norbert Brüggemann, MD[59]; Christine Klein, MD, FEAN[59]; Bastiaan Bloem, MD, PhD[60]

**Coordinators:**
Anisha Singh, BS[23]; Angela Stovall, BS[44]; Julie Festa, BA[1]; Lianne Ramia, BS[1]; Katrina Wakeman, BS[17]; Karen Williams, BA, CCRP[3]; Courtney Blair, MA[18]; Krista Specketer, BS[21]; Diana Willeke[8]; Jennifer Mule, BS[25]; Ella Hilt[26]; Shawnees Peacock, BS[27]; Kori Ribb, RN, BSN, CNRN[28]; Susan Ainscough, BA[30]; Lisbeth Pennente, BA[31]; Julia Brown, BS[32]; Christina Gruenwald, BS, CCRP[32]; Barbara Sommerfeld MSN, RN, CNRN[16]; Farah Kausar, PhD[9]; Alicia Garrido, MD[34]; Deborah Raymond, MS, CGC[14]; Ioana Croitoru[35]; Anne Grete Kristiansen[37]; Helen Mejia Santana, MA[39]; Anjana Singh, BS[20]; Danica Nogo, BS[45]; Shawna Reddie, BA[46]; Samantha Murphy, BS[47]; Lauren O'Brien[48]; Ashwini Ramachandran, MSc[12]; Fnu Madhuri, MS[19]; Daniel Freire, MS[49]; Farah Ismail, MBChB[50]; Raymond James, BS, RN[22]; Tom Osgood, BA, CCRP[51]; Heidi Friedeck, BS[3]; Jenny Frisendahl, BS[52]; Ying Liu, MD[52]; Caitlin Romano, BA[53]; Kelly Clark[24]; Kyle Rizer, BA[54]; Stephanie Carvalho[39]; Sherri Mosovsky, MPH[41]; Farah Sulaiman, MPH[55]; Dora Valent, MS[7]; Raquel Lopes, BSN, MS[29]; Michelle Torreliza, AS[56]; Shira Paz, BS[36]; Victoria Kate Foster[57]; Madita Grümmer[59]; Myrthe Burgler, MA[60]; Sabine van Zundert, MS[60]; Christos Koros, MD, PhD[38]; Jamil Razzaque, MS[58]

**Partners Scientific Advisory Board (Acknowledgement)** ***[Include as applicable to the paper - Obtain current list from MJFF]***

**Abbvie, AcureX, Allergan, Aligning Science Across Parkinson's, Amathus Therapeutics, Avid Radiopharmaceuticals, Bial Biotech, Biohaven, Biogen, BioLegend, Bristol-Myers Squibb, Calico Labs, Celgene, Cerevel, Coave, DaCapo Brainscience, 4D**

**Pharma, Denali, Edmond J. Safra Foundation, Eli Lilly, GE Healthcare, Genentech, GlaxoSmithKline, Golub Capital, Handl Therapeutics, Insitro, Janssen Neuroscience, Lundbeck, Merck, Meso Scale Discovery, Neurocrine Biosciences, Pfizer, Piramal, Prevail Therapeutics, Roche, Sanofi Genzyme, Servier, Takeda, Teva, UCB, VanquaBio, Verily, Voyager Therapeutics, and Yumanity.**

1 Institute for Neurodegenerative Disorders, New Haven, CT
2 23andMe
3 Northwestern University, Chicago, IL
4 University of Iowa, Iowa City, IA
5 VectivBio AG
6 The Michael J. Fox Foundation for Parkinson's Research, New York, NY
7 Innsbruck Medical University, Innsbruck, Austria
8 Paracelsus-Elena Klinik, Kassel, Germany
9 University of California, San Francisco, CA
10 Laboratory of Neuroimaging (LONI), University of Southern California
11 BioRep, Milan, Italy
12 University of Pennsylvania, Philadelphia, PA
13 National Institute on Aging, NIH, Bethesda, MD
14 Mount Sinai Beth Israel, New York, NY
15 Indiana University, Indianapolis, IN
16 Emory University of Medicine, Atlanta, GA
17 Oregon Health and Science University, Portland, OR
18 University of Alabama at Birmingham, Birmingham, AL
19 University of South Florida, Tampa, FL
20 Baylor College of Medicine, Houston, TX
21 University of Washington, Seattle, WA
22 Boston University, Boston, MA
23 University of Rochester, Rochester, NY
24 Banner Research Institute, Sun City, AZ
25 Cleveland Clinic, Cleveland, OH
26 University of Tuebingen, Tuebingen, Germany
27 University of California, San Diego, CA
28 Johns Hopkins University, Baltimore, MD
29 Imperial College of London, London, UK
30 University of Salerno, Salerno, Italy
31 Parkinson's Disease and Movement Disorders Center, Boca Raton, FL
32 University of Cincinnati, Cincinnati, OH
34 Hospital Clinic of Barcelona, Barcelona, Spain
35 Hospital Universitario Donostia, San Sebastian, Spain
36 Tel Aviv Sourasky Medical Center, Tel Aviv, Israel
37 St. Olav's University Hospital, Trondheim, Norway
38 National and Kapodistrian University of Athens, Athens, Greece
39 Columbia University Irving Medical Center, New York, NY
40 Stanford University, Stanford, CA
41 University of Pittsburgh, Pittsburgh, PA
42 Center for Strategy Philanthropy at Milken Institute, Washington D.C.
43 University of Medicine and Dentistry of New Jersey, Piscataway, New Jersey
44 University of Michigan, Ann Arbor, MI
45 Toronto Western Hospital, Toronto, Canada
46 The Ottawa Hospital, Ottawa, Canada
47 Massachusetts General Hospital, Boston, MA
48 University of Kansas Medical Center, Kansas City, KS
49 University of Southern California, Los Angeles, CA
50 Barrow Neurological Institute, Phoenix, AZ
51 Mayo Clinic Arizona, Scottsdale, AZ
52 University of Colorado, Aurora, CO
53 NYU Langone Medical Center, New York, NY
54 University of Florida, Gainesville, FL
55 Montreal Neurological Institute and Hospital/McGill, Montreal, QC, Canada

56 Cleveland Clinic-Las Vegas Lou Ruvo Center for Brain Health, Las Vegas, NV
57 Clinical Ageing Research Unit, Newcastle, UK
58 John Radcliffe Hospital Oxford and Oxford University, Oxford, UK
59 Universität Lübeck, Luebeck, Germany
60 Radboud University, Nijmegen, Netherlands
61 TransThera Consulting
62 Duke University, Durham, NC
